# Deprivation and Exposure to Public Activities during the COVID-19 Pandemic in England and Wales

**DOI:** 10.1101/2021.04.26.21255732

**Authors:** Sarah Beale, Isobel Braithwaite, Annalan M D Navaratnam, Pia Hardelid, Alison Rodger, Anna Aryee, Thomas Byrne, Wing Lam Erica Fong, Ellen Fragaszy, Cyril Geismar, Jana Kovar, Vincent Nguyen, Parth Patel, Madhumita Shrotri, Robert W Aldridge, Andrew Hayward, on behalf of Virus Watch Collaborative, Susan Michie, Linda Wijlaars, Eleni Nastouli, Moira Spyer, Ben Killingley, Ingemar Cox, Vasileios Lampos, Rachel A McKendry, Tao Cheng, Yunzhe Liu, Anne M Johnson, Jo Gibbs, Richard Gilson

## Abstract

**Background:** Differential exposure to public activities and non-household contacts may contribute to stark deprivation-related inequalities in SARS-CoV-2 infection and outcomes, but has not been directly investigated. We set out to investigate whether participants in Virus Watch – a large community cohort study based in England and Wales – reported different levels of exposure to public activities and non-household contacts during the Autumn-Winter phase of the COVID-19 pandemic according to postcode-level socioeconomic deprivation.

**Methods:** Participants (*n*=20120-25228 across surveys) reported their daily activities during three weekly periods in late November 2020, late December 2020, and mid-February 2021. Deprivation was quantified based on participants’ postcode of residence using English or Welsh Indices of Multiple Deprivation quintiles. We used Poisson mixed effect models with robust standard errors to estimate the relationship between deprivation and risk of exposure to public activities during each survey period.

**Results:** Relative to participants in the least deprived areas, participants in the most deprived areas persistently exhibited elevated risk of exposure to vehicle sharing (aRR range across time points 1.73-8.52), public transport (aRR 3.13-5.73), work or education outside of the household (aRR 1.09-1.21), essential shops (aRR 1.09-1.13) and non-household contacts (aRR 1.15-1.19) across multiple survey periods.

**Conclusion:** Differential exposure to essential public activities in deprived communities is likely to contribute to inequalities in infection risk and outcomes during the COVID-19 pandemic. Public health interventions to reduce exposure during essential activities and financial and practical support to enable low-paid workers to stay at home during periods of intense transmission may reduce COVID-related inequalities.

## Introduction

Socioeconomically deprived communities have been disproportionately impacted by the COVID-19 pandemic, experiencing higher rates of infection and mortality than less deprived communities as well as greater social and economic disruption^1–4^.

Public activities - such as working outside the home, visiting shops, or using public transport - may promote the spread of SARS-CoV-2 through contact with potentially infectious individuals and through aerosol transmission^5,6^. Consequently, ‘lockdown’ restrictions - including closure of or access limitations to non-essential shops and services, and recommendations to stay at home - have been a key pandemic response strategy worldwide^7,8^. However, socioeconomic deprivation influences individuals’ ability to stay at home - e.g. as a result of lower ability to work from home and greater reliance on public transport - as well as the level of contact with other people in the workplace. Consequently, differential exposure to public activities may contribute to higher rates of infections, and consequently hospitalisations and deaths from COVID-19, in deprived communities^2,4^.

Empirical investigation into the relationship between daily activities and risk of respiratory infection is limited. Public activities involving potentially poorly-ventilated, high-footfall settings and/or close contact with others - e.g. public transport, visiting cafes or restaurants, shops and supermarkets, venues such as cinemas, theatres and concerts, and social events such as parties - are associated with risk of seasonal respiratory infection^9^. While emerging findings from the COVID-19 pandemic show that measures limiting public mixing are associated with reduced SARS-CoV-2 transmission at the population level^10–12^, data on the nature of individuals’ day-to-day activities are currently limited. Although differential exposure to public activities may influence socioeconomic inequalities in infection and mortality risk, data are lacking regarding how exposure to such activities varies in relation to socio-demographic characteristics including deprivation.

Understanding how deprivation is associated with exposure to public activities is consequently important to ensure that public health and policy responses to COVID-19 and related inequalities are grounded in evidence. We set out to address this gap using data on public activities and non-household contacts collected through the Virus Watch cohort study^13^.

## Methods

### Survey Procedure

We used data from three consecutive survey waves of the Virus Watch household cohort study^13^. Households were recruited via post, social media, SMS messages, and personalised letters disseminated by General Practices. Eligibility criteria were: residence in England or Wales, informed consent or assent provided by all household members, internet access and an email address, at least one household member able to complete surveys in English, and household size between 0-6 household members (due to survey infrastructure limitations). Virus Watch study procedures are described in detail in the study protocol^13^ and include completion of monthly questionnaires into pandemic-relevant demographic, psychosocial/behavioral and health-related factors from which the current data were drawn.

Participants were prompted on 01 December 2020, 04 January 2021, and 17 February 2021 to complete an online questionnaire regarding their social activities and contacts during the preceding week. These survey weeks corresponded to key timepoints in terms of epidemic waves and/or government legislation regarding public activities in England and Wales. The first survey (covering 24 Nov 2020 - 01 Dec 2020) corresponded to the final week of the second English national lockdown^14^, and the early stages of a sharp rise in COVID-19 cases and the emergence of the Lineage B.1.1.7 variant. The second survey week (23 Dec 2020 - 27 Dec 2020) corresponded to the December holiday period, during which there was notable variation in rules around social mixing across regions that was altered in the run-up to the holiday period due to sharp increases in COVID-19 cases in some regions. Indoor mixing with non-household members was not allowed in London, the South East or East of England, while indoor mixing on the 25^th^ December was allowed with a maximum of three other households across other English regions^15^ and with two other households in Wales^16^. The third survey week (09 Feb 2021 - 16 Feb 2021) occurred during the third national lockdown for both England and Wales^16,17^ – a period of deceleration in reported cases nationally.

Survey respondents reported the days they undertook a range of activities during each period (see Outcomes below) and their number of non-household/support bubble close contacts (‘face-to-face contact with someone less than a metre away, even if a face-covering or mask was worn, or within 2 metres for 15 minutes or more’^18^). The wording of questions in the second and third surveys was edited to specifically refer to contact with non-household or support bubble members (vs non-household members) for clarity. Survey data were extracted on the 25 Feb 2021.

### Exposure

The exposure of interest, deprivation, was measured at small local area-level based on the Ministry for Housing and Local Government (in England) and Welsh Government (in Wales) Index of Multiple Deprivation (IMD) quintiles (1=most deprived, 5= least deprived). Participants provided household postcodes on study registration, which were used to derive IMD quintiles based on linkage with the May 2020 ONS Postcode Lookup file. Consequently, only survey respondents who provided a valid postcode at baseline were included in these analyses.

### Outcome

The following activities were classified as binary outcomes of interest (yes/no during given period): driving or riding in a car/taxi with a non-household member, taking public transport (underground trains, overground trains, buses, or trams), going to work or education outside the household, social/entertainment activities (defined as any of: attending the theatre, cinema, concert or sports event; eating in a restaurant, café or canteen; going to a bar, pub or club; going to a party), going to essential shops, going to non-essential shops or personal care services, and close contact with one or more non-household/support bubble members. In the first and second surveys, exposure to car/taxis was asked as a single item; in the third survey, this item was disaggregated into separate car and taxi exposures.

### Covariates

Age, sex, and geographic region were considered relevant a-priori potential confounders due to plausible relationships with both IMD and activities. Age and sex were derived from participants’ responses to demographic questions at study baseline. Age was classified as child (0-15), adult (16-64), and older adult (65+). Region was derived from linking participants’ postcode to ONS national region using the May 2020 National Statistics Postcode Lookup file. For the current analyses, regions were classified into the following three categories based on differing activity-related legislation and rates of SARS-CoV-2: London/South-East/ East of England (initial Tier 4 regions), Wales, and other regions.

### Statistical Analyses

To assess the age-, sex-, and region-adjusted risk of reporting each activity by IMD quintile, we used Poisson mixed-effect models with robust standard errors^19^ using the *mepoisson* command in Stata version 16. All available data were entered into the models; data were complete for IMD and region and missing data were limited across time points for age group (range across time 0.29%-0.35%, *n=*59-84) and sex (range 1.06%-1.48%, *n=*217-310). The least deprived quintile (IMD 5) was used as the reference category. We included a random term to account for household-level clustering. We applied the Benjamini-Hochberg Procedure (false discovery rate = 0.05) to correct for multiple testing.

We performed a sensitivity analysis stratifying the relationship between IMD and attending work/education settings by age (child <16 years vs adult ≥16 years) to account for potential effect modification, as the impact of legislation around school openings may have had a more consistent effect on children across IMD quintiles than legislation around work/higher education for adults. As attendance of work/education settings is likely to influence non-household contacts, we also stratified the relationship between IMD and non-household contacts by age. For adults, we also performed a further sensitivity analysis for these outcomes controlling for presence of children (<16) in the household. This was considered a potential confounder due to the negative association between maternal education (likely associated with small area-level deprivation) and likelihood of having children^20^, along with the likely influence of COVID-related school closures on the working patterns of parents and carers.

### Ethics and Consent

The Virus Watch study was approved by the Hampstead NHS Health Research Authority Ethics Committee: 20/HRA/2320, and conformed to the ethical standards set out in the Declaration of Helsinki. All members of participating households provided informed consent for themselves and, where relevant, for dependent children.

## Results

Table 1 reports the characteristics of the full Virus Watch cohort as of 25 Feb 2021 (*n*=46539 individuals from 22556 households) and of respondents to each activity survey who provided a valid postcode at baseline (*n*=20120-25228).

**Table 1.**
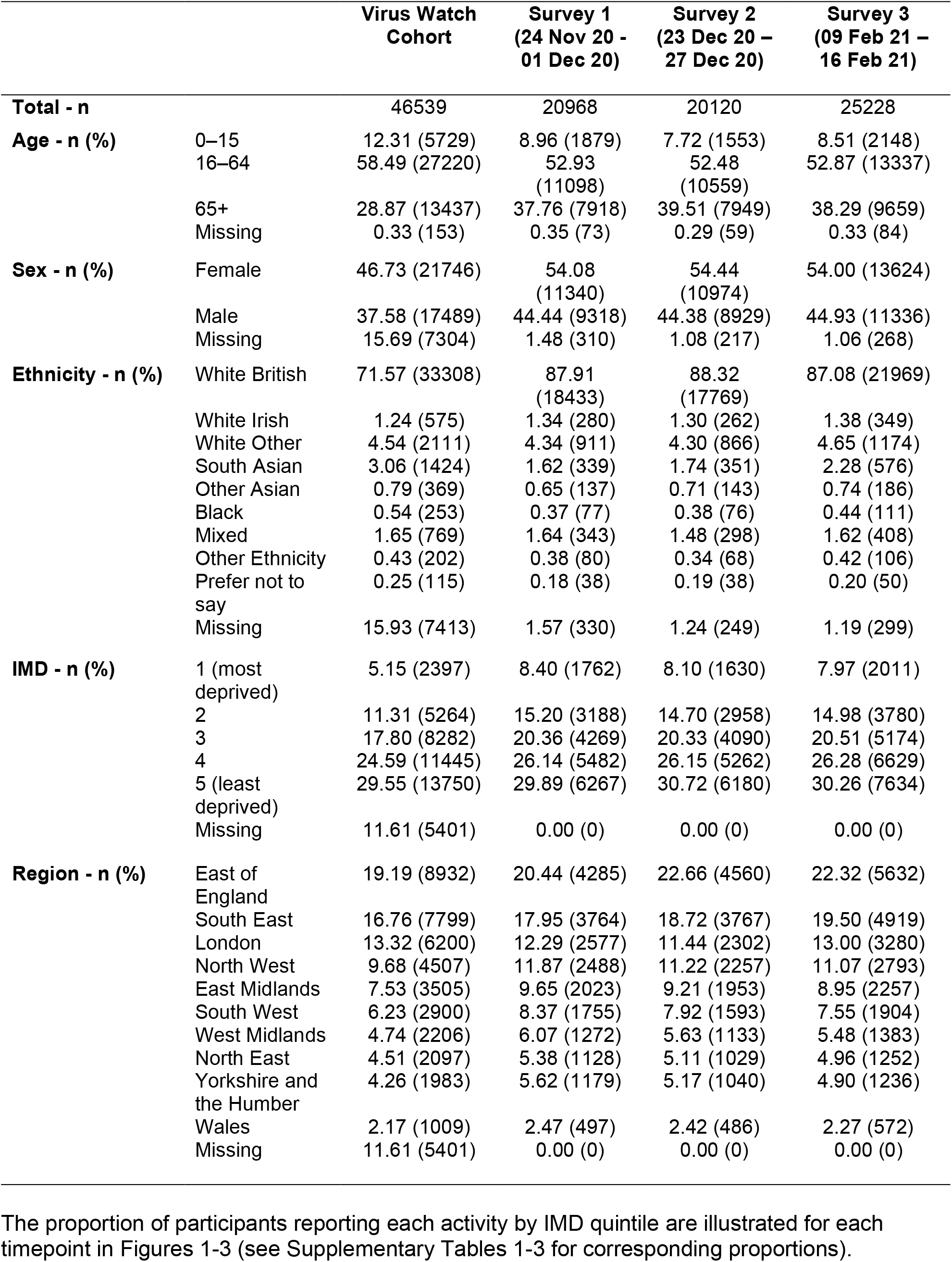
Demographic Features of Survey Respondents.

|  |  | **Virus Watch Cohort** | **Survey 1**<br>**(24 Nov 20 - 01 Dec 20)** | **Survey 2**<br>**(23 Dec 20 - 27 Dec 20)** | **Survey 3**<br>**(09 Feb 21 - 16 Feb 21)** |
| --- | --- | --- | --- | --- | --- |
| **Total - n** |  | 46539 | 20968 | 20120 | 25228 |
| **Age - n (%)** | 0–15 | 12.31 (5729) | 8.96 (1879) | 7.72 (1553) | 8.51 (2148) |
|  | 16–64 | 58.49 (27220) | 52.93 (11098) | 52.48 (10559) | 52.87 (13337) |
|  | 65+ | 28.87 (13437) | 37.76 (7918) | 39.51 (7949) | 38.29 (9659) |
|  | Missing | 0.33 (153) | 0.35 (73) | 0.29 (59) | 0.33 (84) |
| **Sex - n (%)** | Female | 46.73 (21746) | 54.08 (11340) | 54.44 (10974) | 54.00 (13624) |
|  | Male | 37.58 (17489) | 44.44 (9318) | 44.38 (8929) | 44.93 (11336) |
|  | Missing | 15.69 (7304) | 1.48 (310) | 1.08 (217) | 1.06 (268) |
| **Ethnicity - n (%)** | White British | 71.57 (33308) | 87.91 (18433) | 88.32 (17769) | 87.08 (21969) |
|  | White Irish | 1.24 (575) | 1.34 (280) | 1.30 (262) | 1.38 (349) |
|  | White Other | 4.54 (2111) | 4.34 (911) | 4.30 (866) | 4.65 (1174) |
|  | South Asian | 3.06 (1424) | 1.62 (339) | 1.74 (351) | 2.28 (576) |
|  | Other Asian | 0.79 (369) | 0.65 (137) | 0.71 (143) | 0.74 (186) |
|  | Black | 0.54 (253) | 0.37 (77) | 0.38 (76) | 0.44 (111) |
|  | Mixed | 1.65 (769) | 1.64 (343) | 1.48 (298) | 1.62 (408) |
|  | Other Ethnicity | 0.43 (202) | 0.38 (80) | 0.34 (68) | 0.42 (106) |
|  | Prefer not to say | 0.25 (115) | 0.18 (38) | 0.19 (38) | 0.20 (50) |
|  | Missing | 15.93 (7413) | 1.57 (330) | 1.24 (249) | 1.19 (299) |
| **IMD - n (%)** | 1 (most deprived) | 5.15 (2397) | 8.40 (1762) | 8.10 (1630) | 7.97 (2011) |
|  | 2 | 11.31 (5264) | 15.20 (3188) | 14.70 (2958) | 14.98 (3780) |
|  | 3 | 17.80 (8282) | 20.36 (4269) | 20.33 (4090) | 20.51 (5174) |
|  | 4 | 24.59 (11445) | 26.14 (5482) | 26.15 (5262) | 26.28 (6629) |
|  | 5 (least deprived) | 29.55 (13750) | 29.89 (6267) | 30.72 (6180) | 30.26 (7634) |
|  | Missing | 11.61 (5401) | 0.00 (0) | 0.00 (0) | 0.00 (0) |
| **Region - n (%)** | East of England | 19.19 (8932) | 20.44 (4285) | 22.66 (4560) | 22.32 (5632) |
|  | South East | 16.76 (7799) | 17.95 (3764) | 18.72 (3767) | 19.50 (4919) |
|  | London | 13.32 (6200) | 12.29 (2577) | 11.44 (2302) | 13.00 (3280) |
|  | North West | 9.68 (4507) | 11.87 (2488) | 11.22 (2257) | 11.07 (2793) |
|  | East Midlands | 7.53 (3505) | 9.65 (2023) | 9.21 (1953) | 8.95 (2257) |
|  | South West | 6.23 (2900) | 8.37 (1755) | 7.92 (1593) | 7.55 (1904) |
|  | West Midlands | 4.74 (2206) | 6.07 (1272) | 5.63 (1133) | 5.48 (1383) |
|  | North East | 4.51 (2097) | 5.38 (1128) | 5.11 (1029) | 4.96 (1252) |
|  | Yorkshire and the Humber | 4.26 (1983) | 5.62 (1179) | 5.17 (1040) | 4.90 (1236) |
|  | Wales | 2.17 (1009) | 2.47 (497) | 2.42 (486) | 2.27 (572) |
|  | Missing | 11.61 (5401) | 0.00 (0) | 0.00 (0) | 0.00 (0) |

**Figure 1.**
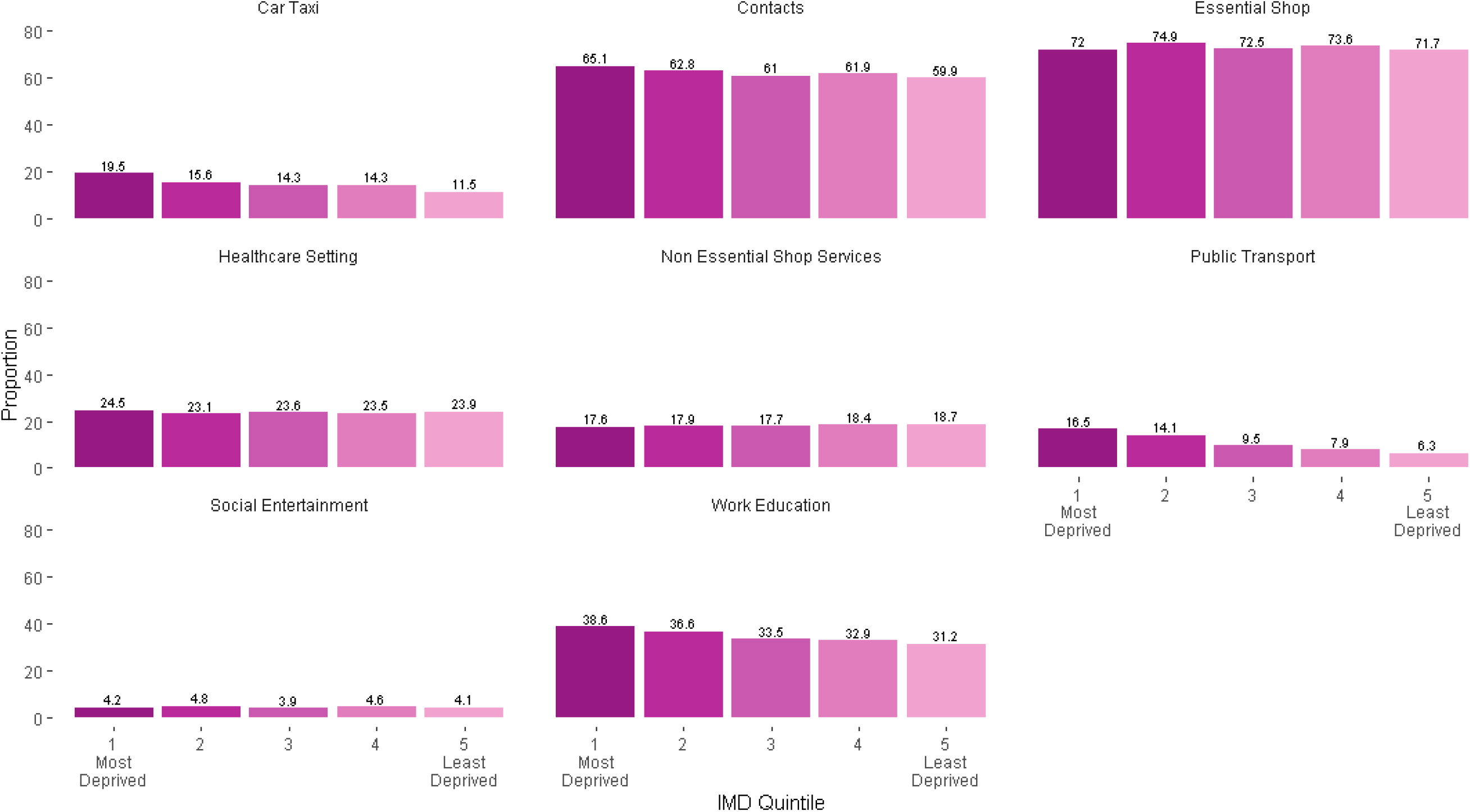
Proportion of Participants Reporting Public Activities and Non-Household Contacts by IMD Quintile (24 Nov 20 – 01 Dec 20)

**Figure 2.**
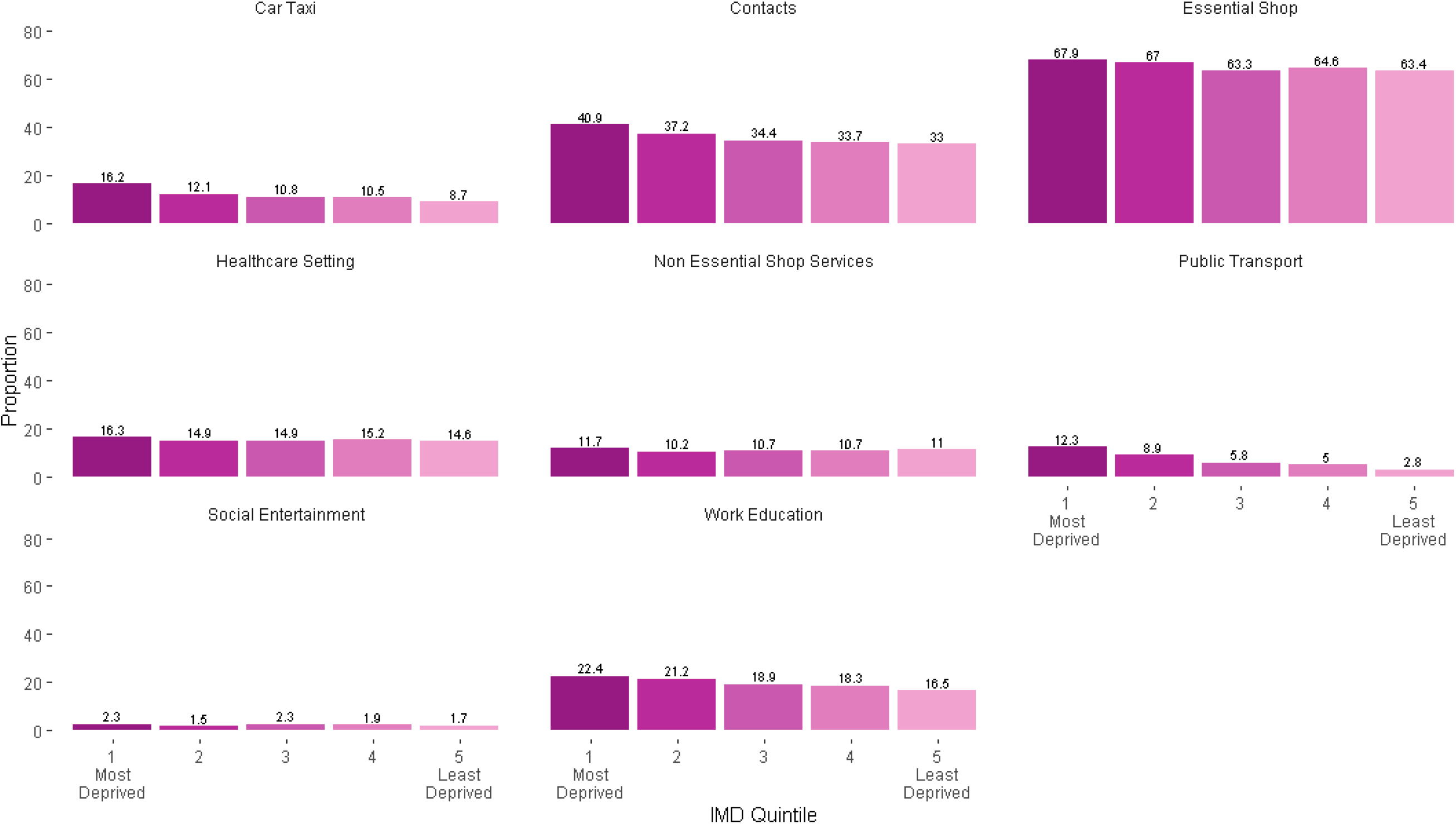
Proportion of Participants Reporting Public Activities and Non-Household Contacts by IMD Quintile (23 Dec 20 - 27 Dec 20)

**Figure 3.**
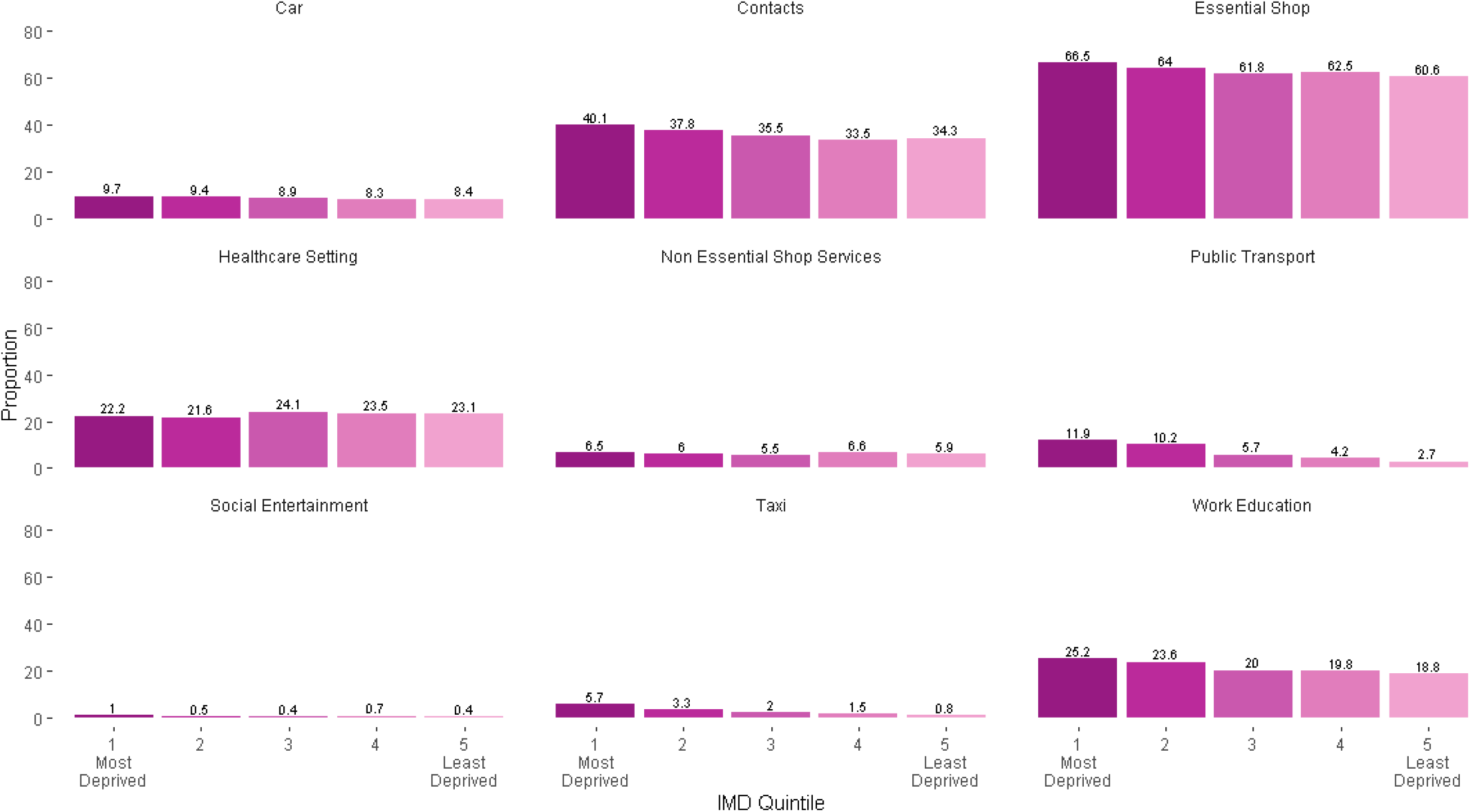
Proportion of Participants Reporting Public Activities and Non-Household Contacts by IMD Quintile (09 Feb 21 - 16 Feb 21)

Poisson mixed models for the first survey period (24 Nov 2020 - 01 Dec 2020) (Table 2 and Supplementary Figure 1a/b) indicated that – compared to the least deprived group – participants in all other IMD quintiles had elevated risk following multiple comparison correction for vehicle-sharing (aRR range: 1.22 [1.08-1.38] – 1.73 [1.50-1.99]) and for attending work or education outside the home (aRR range: 1.07 [1.01-1.12] – 1.12 [1.03-1.18]). When stratified by age, adults - but not children - in IMD quintiles 1 and 2 demonstrated elevated risk of exposure to work or education settings (Supplementary Table 4), and adults in IMD 1-3 demonstrated elevated risk after controlling for presence of children in the household (Supplementary Table 6). Participants in IMD quintiles 1-3 also had an elevated risk of exposure to public transport (aRR range: 1.41 [1.21-1.65] – 3.13 [2.63-3.70]). Confidence intervals indicated that the two most deprived quintiles had elevated risk of exposure to public transport compared to all other quintiles. Relative to the least deprived IMD quintile, the second most deprived quintile had greater risk of visiting essential shops (aRR: 1.07 [1.04-1.10]). No differences emerged by IMD for social and entertainment activities, visiting non-essential shops, or healthcare settings. While no difference emerged in the main analyses for exposure to non-household contacts, age-stratified analysis (Supplementary Table 5) indicated elevated risk for adults in IMD 1 and 2 (aRR respectively: 1.11 [1.06-1.18] and 1.07 [1.02-1.12]), which persisted after controlling for the presence of children in the household (Supplementary Table 6).

**Table 2.** Risk Ratios for Public Activities and Non-Household Contacts by IMD Quintile.

| Survey Period | IMD | Car Sharing or Taxi | Public Transport | Work or Education | Social and Entertainment Activities | Essential Shops | Non-Essential Shops and Services | Healthcare SettingS | Non-Household Contacts |
| --- | --- | --- | --- | --- | --- | --- | --- | --- | --- |
| 24 Nov 20 - 01 Dec 20 | 1 | 1.73*<br>[1.50,1.99] | 3.13*<br>[2.63,3.70] | 1.09*<br>[1.01,1.17] | 1.10<br>[0.79,1.53] | 1.04<br>[1.00,1.08] | 0.96<br>[0.83,1.10] | 1.11<br>[1.00,1.23] | 1.06<br>[1.01,1.12] |
|  | 2 | 1.37*<br>[1.21,1.55] | 2.27*<br>[1.96,2.63] | 1.12*<br>[1.03,1.18] | 1.22<br>[0.95,1.38] | 1.07*<br>[1.04,1.10] | 0.98<br>[0.87,1.09] | 1.00<br>[0.92,1.09] | 1.03<br>[0.99,1.07] |
|  | 3 | 1.22*<br>[1.08,1.38] | 1.41*<br>[1.21,1.65] | 1.07*<br>[1.01,1.13] | 0.99<br>[0.77,1.27] | 1.01<br>[0.99,1.04] | 0.97<br>[0.87,1.07] | 0.99<br>[0.92,1.07] | 1.01<br>[0.97,1.05] |
|  | 4 | 1.23*<br>[1.10,1.38] | 1.28<br>[1.10,1.49] | 1.07*<br>[1.01,1.12] | 1.20<br>[0.96,1.30] | 1.03<br>[1.00,1.05] | 0.98<br>[0.90,1.08] | 0.99<br>[0.92,1.06] | 1.03<br>[1.00,1.07] |
|  | 5 | REF | REF | REF | REF | REF | REF | REF | REF |
| 23 Dec 20 - 27 Dec 20) | 1 | 1.97*<br>[1.65,2.35] | 5.53*<br>[4.37,7.00] | 1.17*<br>[1.04,1.31] | 1.43<br>[0.87,2.34] | 1.09*<br>[1.04,1.14] | 0.96<br>[0.79,1.15] | 1.17<br>[1.02,1.35] | 1.19*<br>[1.09,1.29] |
|  | 2 | 1.45*<br>[1.24,1.70] | 3.41*<br>[2.64,4.24] | 1.20*<br>[1.09,1.33] | 1.00<br>[0.64,1.55] | 1.07*<br>[1.04,1.11] | 0.92<br>[0.79,1.07] | 1.06<br>[0.94,1.19] | 1.11*<br>[1.03,1.20] |
|  | 3 | 1.24*<br>[1.07,1.45] | 2.03*<br>[1.61,2.56] | 1.14*<br>[1.04,1.25] | 1.34<br>[0.90,1.99] | 1.01<br>[0.98,1.04] | 1.05<br>[0.91,1.21] | 1.03<br>[0.93,1.15] | 1.05<br>[0.98,1.12] |
|  | 4 | 1.23*<br>[1.06,1.42] | 1.80*<br>[1.44,2.25] | 1.11<br>[1.02,1.22] | 1.10<br>[0.75,1.60] | 1.02<br>[0.99,1.05] | 0.96<br>[0.84,1.10] | 1.04<br>[0.94,1.14] | 1.02<br>[0.96,1.09] |
|  | 5 | REF | REF | REF | REF | REF | REF | REF | REF |
| 09 Feb 21 -<br>16 Feb 21 | 1 | Car: 1.13<br>[0.94,1.37];<br>Taxi: **8.52\***<br>**[5.96,12.17]** | **5.73\***<br>**[4.65,7.05]** | **1.21\***<br>**[1.10,1.34]** | **2.64\***<br>**[1.41,4.93]** | **1.13\***<br>**[1.08,1.17]** | 1.14<br>[0.90,1.44] | 1.04<br>[0.94,1.15] | **1.15\***<br>**[1.06,1.24]** |
|  | 2 | Car: 1.14<br>[0.98,1.33];<br>Taxi: **4.42\***<br>**[3.14,6.20]** | **4.05\***<br>**[3.36,4.89]** | **1.20\***<br>**[1.11,1.31]** | 1.22<br>[0.67,2.20] | **1.08\***<br>**[1.04,1.12]** | 1.07<br>[0.88,1.30] | 0.99<br>[0.91,1.07] | 1.09<br>[1.02,1.17] |
|  | 3 | Car: 1.09<br>[0.95,1.26];<br>Taxi: **2.49\***<br>**[1.75,3.53]** | **2.06\***<br>**[1.69,2.51]** | 1.08<br>[0.99,1.17] | 1.05<br>[0.60,1.84] | 1.03<br>[1.00,1.06] | 0.97<br>[0.82,1.16] | 1.06<br>[0.98,1.13] | 1.03<br>[0.97,1.10] |
|  | 4 | Car: 0.98<br>[0.86,1.13];<br>Taxi: 1.75<br>[1.21,2.52] | **1.59\***<br>**[1.29,1.95]** | 1.06<br>[0.99,1.15] | 1.32<br>[0.79,2.19] | 1.03<br>[1.00,1.06] | 1.13<br>[0.97,1.33] | 1.02<br>[0.95,1.09] | 0.98<br>[0.92,1.04] |
|  | 5 | REF | REF | REF | REF | REF | REF | REF | REF |

**Note:** IMD = Indices of Multiple Deprivation (1 = most deprived; 5 = least deprived); 95% confidence intervals in parentheses
\* **Bonferroni-Hochberg adjusted $p < 0.05$**

In the second survey period (23 Dec 2020 - 27 Dec 2020) (Table 2 and Supplementary Figure 2a/b) all investigated IMD groups had elevated risk of vehicle-sharing (aRR range: 1.24 [1.07-1.45] – 1.97 [1.65-2.35]) and public transport use (aRR range: 1.80 [1.44-2.45] – 5.53 [4.37-7.00]) compared to the least deprived group. As above, confidence intervals for IMD 1-2 indicated elevated risk for public transport compared to all other quintiles. IMD quintiles 1-3 also had an elevated risk of attending work or education outside the home (aRR range: 1.14 [1.04-1.25] – 1.20 [1.09-1.33]); when stratified by age, this finding was consistent for adults only (Supplementary Table 4) and remained following adjustment for presence of children in the household (Supplementary Table 6). IMD quintiles 1 and 2 reported elevated risk of visiting an essential shop (aRR respectively: 1.09 [1.04-1.14] and 1.07 [1.04-1.11]). These groups also demonstrated elevated risk of exposure to non-household contacts (aRR respectively: 1.19 [1.09-1.29] and 1.11 [1.03-1.20]), which was present only for adults after age-stratification (Supplementary Table 5) and remained following adjustment for children in the household (Supplementary Table 6). Again, no differences were identified for social and entertainment activities, visiting non-essential shops, or healthcare settings.

Consistent with previous timepoints, all investigated IMD groups had elevated risk of reporting public transport use during the third survey period (09 Feb 2021-16 Feb 2021) (Table 2 and Supplementary Figure 3a/b) compared to the least deprived group (aRR range: 1.59 [1.29-1.95] – 5.73 [4.65-7.05]). Confidence intervals for IMD 1 and 2 again indicated elevated risk of exposure to public transport compared to all other IMD quintiles. Car-sharing versus taxi use were disaggregated in this survey, and IMD 1-3 had elevated risk of taxi use compared to the least deprived group (aRR range 2.49 [1.75-3.53] - 8.52 [4.65-7.05]). No difference was found by IMD for car sharing, which was less-commonly reported overall (supplementary Table 2). Similar to previous timepoints, IMD 1-2 had elevated risk of exposure to essential shops (aRR respectively 1.13 [1.08-1.17] and 1.08 [1.04-1.12]) and to workplace or education settings outside the home (aRR respectively 1.21 [1.10-1.34] and 1.20 [1.11-1.31]). Again, age-stratification (Supplementary Table 4) indicated no difference in risk of exposure to work/education settings for children, while adults in IMD 1 and 2 demonstrated elevated risk. Adults in IMD 1-3 demonstrated excess risk of exposure to these settings following adjustment for children in the household. IMD 1 also had elevated risk of exposure to non-household contacts (aRR: 1.15 [1.06-1.24]); excess risk was demonstrated by adults in IMD 1 and 2 following age stratification (Supplementary Table 5), and IMD 1-3 following adjustment for presence of children in the household (Supplementary Table 6). For the first time across measured timepoints, participants in IMD 1 also demonstrated excess risk of reporting social and entertainment activities (aRR: 2.64 [1.41-4.93]). No differences were found by IMD for visiting non-essential shops or healthcare settings.

## Discussion

Our findings suggest that differences in essential daily activities – such as using public transport, attending work/education settings outside the home, and visiting essential shops – likely contribute to elevated risk of COVID-19 infection and mortality in deprived communities. Patterns of differential exposure to essential activities were consistent across the three survey periods. Adults - but not children - consistently demonstrated differential exposure to workplace/education settings and non-household contacts, likely reflecting the effects of legislation around school openings for children. We found limited evidence of deprivation-related differences for activities more reflective of individual choice, with no differences in visiting non-essential shops or services and elevated risk of social and entertainment activities in the most deprived group at one time-point only (February 2021).

This is the first study to investigate the relationship between deprivation and exposure to specific daily activities during the COVID-19 pandemic. The Virus Watch cohort comprises households from across England and Wales with considerable diversity in age, ethnicity, and area-level deprivation. However, survey respondents were not demographically representative of the population. Self-reported activities may have been affected by recall and social desirability biases, though the latter may have been minimised by online survey delivery. A further limitation was the inability to quantify infection risk associated with each activity due to insufficient data. This will be the focus of future work when additional virological and serological outcome data are available. Quantifying the number and intensity of contacts per setting and any risk mitigation strategies were also beyond the scope of these surveys. Despite relevant age-stratified analysis, we were not able to directly distinguish between workplace and education settings in the current study. IMD is also an area-level measure and may not always reflect individuals’ socioeconomic position. Further investigation into the influence of individual-level indicators of socio-economic position including occupation and education is warranted, as is investigation into the interrelationship between deprivation, ethnicity, and infection risk. Investigating workplace attendance by occupation is also an important area for further investigation given differential exposure to workplace/education settings for adults.

In the UK, lockdown and social distancing measures appear to have reduced contacts and mobility in public locations compared to pre-pandemic levels^21–23^; however, the current findings suggest that deprivation-related differences in exposure to essential public activities are consistently present during periods of stringent regulations. These findings are consistent with a USA-based study^24^ that constructed granular spatio-temporal mobility networks based on mobile phone data for 98 million people across ∼57,000 neighbourhoods. Integrating these networks within a metapopulation susceptible-exposed-infectious-recovered model allowed an accurate fit to observed case trajectories and predicted ethnicity- and deprivation-related risk gradients. This study found that disadvantaged groups’ mobility did not reduce as sharply as people living in majority-white and higher-income areas, and that the locations that they visited were typically smaller and more crowded.

Interpretation of activity and contact patterns reported in the current study should take into account changes in the wider context at these times. The lockdown restrictions that were in place during the November and February surveys were similar in England; key differences being the closure of primary and secondary schools and increased fines in January and February^14,17^. The English policy context surrounding the December survey differed substantially: different ‘tier’-based restrictions were in place across England, and rules for contact on December 25 (Christmas Day) were relaxed only in some regions. No indoor mixing with non-household members was allowed in London, the South East or East of England; other English regions were allowed meeting a maximum of three households^15^. In Wales, the late November survey fell two weeks after the 17-day national ‘firebreak’ lockdown ended, and a further Welsh lockdown started on December 20^16^, with rules relaxed to allow mixing with up to two other households on December 25 only^16^. In February, Wales was again under lockdown. Differential exposure to essential activities and non-household contacts was consistently observed across survey periods. Consequently, deprivation-related inequalities in infection and mortality risk driven by essential public activities may persist or become exacerbated during periods of stringent lockdown restrictions. Measuring differential activities during periods of less intensive or no restrictions on social mixing is consequently warranted.

The current findings suggest that interventions to reduce SARS-CoV-2 exposure on public transport, in essential shops, and in workplace and education settings where in-person attendance is required may reduce inequalities in infection risk. Deprivation-related differences in exposure to these essential activities are likely to reflect structural factors that constrain individual choice, such as car ownership, ability to work from home and disposable income. Providing greater financial and practical support to facilitate increased uptake of testing and adherence to isolation when required may also prevent mortality and reduce inequalities^25^. Interventions and public health communications targeting activities that do not consistently differ between social groups over time, such as fines for attending social activities, conversely appear unlikely to reduce COVID-related inequalities.

## Data Availability

We aim to share aggregate data from this project on our website and via a "Findings so far" section on our website - https://ucl-virus-watch.net/. We will also be sharing individual record level data with personal identifiers removed on a research data sharing service such as the Office of National Statistics Secure Research Service. In sharing the data we will work within the principles set out in the UKRI Guidance on best practice in the management of research data. Access to use of the data whilst research is being conducted will be managed by the Chief Investigators (ACH and RWA) in accordance with the principles set out in the UKRI guidance on best practice in the management of research data. It is the intention that the data arising from this research will initially be collected, cleaned and validated by the UCL research team and once this has been completed will be shared for wider use. We aim to make subsets of the data more rapidly available both on our study website and via the public facing dashboard during the ongoing phase of data collection. In line with Principle 5 of the UKRI guidance on best practice in the management of research data, we plan to release data in batches as they become available or as updated results are published. Individual record data linked using NHS Digital will not be shared, only aggregated results. HES and mortality data may be obtained from a third party and are not publicly available. These data are owned by a third party and can be accessed by researchers applying to the Health and Social Care Information Centre for England. We will put analysis code on publicly available repositories to enable their reuse.

https://ucl-virus-watch.net/

## References

1. Marmot M, Allen J. COVID-19: exposing and amplifying inequalities. J Epidemiol Community Health 2020;74(9):681–2.

2. Office for National Statistics. Deaths involving COVID-19 by local area and socioeconomic deprivation: deaths occurring between 1 March and 31 July 2020. https://www.ons.gov.uk/peoplepopulationandcommunity/birthsdeathsandmarriages/deaths/bulletins/deathsinvolvingcovid19bylocalareasanddeprivation/deathsoccurringbetween1marchand31july2020 [Accessed 18/04/2021].

3. Patel AP, Paranjpe MD, Kathiresan NP, et al. Race, socioeconomic deprivation, and hospitalization for COVID-19 in English participants of a national biobank. International Journal for Equity in Health 2020;19(1):1–4.

4. Public Health England. Disparities in the risk and outcomes of COVID-19. https://assets.publishing.service.gov.uk/government/uploads/system/uploads/attachment_data/file/908434/Disparities_in_the_risk_and_outcomes_of_COVID_August_2020_update.pdf [Accessed 18/04/2021].

5. Jayaweera M, Perera H, Gunawardana B, et al. Transmission of COVID-19 virus by droplets and aerosols: A critical review on the unresolved dichotomy. Environmental Research 2020; 109819.

6. Meyerowitz EA, Richterman A, Gandhi RT, et al. Transmission of SARS-CoV-2: a review of viral, host, and environmental factors. Annals of Internal Medicine 2020.

7. Cabinet Office. Staying at home and away from others (social distancing). https://www.gov.uk/government/publications/full-guidance-on-staying-at-home-and-away-from-others/full-guidance-on-staying-at-home-and-away-from-others [Accessed 18/04/2021].

8. World Health Organisation. Coronavirus disease 2019 (COVID-19) Advice for the Public. https://www.who.int/emergencies/diseases/novel-coronavirus-2019/advice-for-public [Accessed 18/04/2021]

9. Hayward AC, Beale S, Johnson AM, et al. Public activities preceding the onset of acute respiratory infection syndromes in adults in England-implications for the use of social distancing to control pandemic respiratory infections. Wellcome Open Research 2020;5.

10. Brauner JM, Mindermann S, Sharma M, et al. Inferring the effectiveness of government interventions against COVID-19. Science 2020.

11. Dehning J, Zierenberg J, Spitzner FP, et al. Inferring change points in the spread of COVID- 19 reveals the effectiveness of interventions. Science 2020;369(6500).

12. Ingelbeen, B, Peckeu, L, Laga, M, et al. Reducing contacts to stop SARS-CoV-2 transmission during the second pandemic wave in Brussels, Belgium, August to November 2020. Eurosurveillance 2021; 2100065.

13. Hayward A, Fragaszy E, Kovar J, et al. Risk factors, symptom reporting, healthcare-seeking behaviour and adherence to public health guidance: protocol for Virus Watch, a prospective community cohort study. medRxiv 2020.

14. Cabinet Office. Coronavirus (COVID-19): New National Restrictions from 5 November; https://www.gov.uk/guidance/new-national-restrictions-from-5-november [Accessed 18/04/2021].

15. Cabinet Office. Coronavirus (COVID-19): Guidance for the Christmas Period. [Accessed 18/04/2021].

16. Welsh Government. Coronavirus (COVID-19). https://gov.wales/coronavirus [Accessed 18/04/2021].

17. Cabinet Office. Coronavirus (COVID-19): National lockdown: Stay at Home. https://www.gov.uk/guidance/national-lockdown-stay-at-home [Accessed 18/04/2021].

18. Department of Health and Social Care. NHS Test and Trace: what to do if you are contacted: https://www.gov.uk/guidance/nhs-test-and-trace-how-it-works [Accessed 18/04/2021].

19. Zou G. A modified poisson regression approach to prospective studies with binary data. American Journal of Epidemiology 2004; 159(7):702–6.

20. Berrington A. Childlessness in the UK. In Childlessness in Europe: Contexts, causes, and consequences 2017 (pp. 57–76). Springer, Cham.

21. Jarvis CI, Gimma A, van Zandvoort K, et al. CoMix study. Social contact survey in the UK. https://cmmid.github.io/topics/covid19/comix-reports.html [Accessed 18/04/2021].

22. Apple. Mobility Trends. https://www.apple.com/covid19/mobility. [Accessed 18/04/2021].

23. Google. Community mobility reports. https://www.google.com/covid19/mobility/index.html?hl=en. [Accessed 18/04/2021].

24. Chang S, Pierson E, Koh PW, et al. Mobility network models of COVID-19 explain inequities and inform reopening. Nature 2021;589(7840):82–7.

25. Scientific Pandemic Insights Group on Behaviours (SPI-B). The impact of financial and other targeted support on rates of self-isolation or quarantine. https://assets.publishing.service.gov.uk/government/uploads/system/uploads/attachment_data/file/925133/S0759_SPI-BThe_impact_of_financial_and_other_targeted_support_on_rates_of_self-isolation_or_quarantine_.pdf [Accessed 18/04/2021].

