## Supplementary Materials for "Deprivation and Exposure to Public Activities during the COVID-19 Pandemic in England and Wales"

### Supplementary Material

#### *Supplementary Table 1.*

**Proportion of participants reporting each activity and total number of respondents by deprivation quintile (24 Nov 20 – 01 Dec 20)**

| **IMD Quintile** | **Car Sharing or Taxi** | | **Public Transport** | | **Work or Education** | | **Social/ Entertainment** | | **Essential Shops** | | **Non-Essential Shops/Services** | | **Healthcare Setting** | | **Non-Household Contacts** | |
|---|---|---|---|---|---|---|---|---|---|---|---|---|---|---|---|---|
| | Yes % (*n*) | total *n* | Yes % (*n*) | total *n* | Yes % (*n*) | total *n* | Yes % (*n*) | total *n* | Yes % (*n*) | total *n* | Yes % (*n*) | total *n* | Yes % (*n*) | total *n* | Yes % (*n*) | total *n* |
| **1 (Most Deprived)** | 19.52 (344) | 1762 | 16.52 (291) | 1762 | 38.57 (680) | 1763 | 4.20 (74) | 1762 | 72.04 (1270) | 1763 | 17.58 (310) | 1763 | 24.52 (432) | 1762 | 65.06 (1145) | 1760 |
| **2** | 15.63 (498) | 3188 | 14.20 (450) | 3186 | 36.59 (1166) | 3187 | 4.84 (154) | 3184 | 74.91 (2389) | 3189 | 17.86 (569) | 3185 | 23.05 (734) | 3184 | 62.77 (2003) | 3191 |
| **3** | 14.34 (612) | 4269 | 9.49 (405) | 4267 | 33.52 (1431) | 4269 | 3.89 (166) | 4267 | 72.52 (3096) | 4269 | 17.71 (756) | 4268 | 23.63 (1008) | 4266 | 60.97 (2599) | 4263 |
| **4** | 14.30 (784) | 5482 | 7.89 (432) | 5478 | 32.88 (1801) | 5478 | 4.56 (250) | 5477 | 73.64 (4034) | 5478 | 18.39 (1007) | 5476 | 23.52 (1289) | 5480 | 61.88 (3391) | 5480 |
| **5 (Least Deprived)** | 11.54 (723) | 6267 | 6.27 (393) | 6266 | 31.16 (1952) | 6268 | 4.12 (258) | 6268 | 71.68 (4493) | 6268 | 18.69 (1171) | 6266 | 23.92 (1499) | 6268 | 59.85 (3748) | 6262 |
| **Total** | 14.12 (2961) | 20968 | 9.40 (1971) | 20959 | 33.54 (7031) | 20965 | 4.30 (902) | 20958 | 72.89 (15282) | 20967 | 18.19 (3813) | 20958 | 23.67 (4962) | 20960 | 61.49 (12886) | 20956 |

**Note:** IMD = Indices of Multiple Deprivation; total respondents for each question reported separately due to varying frequencies across questions

*Supplementary Table 2.*

**Proportion of participants reporting each activity and total number of respondents by deprivation quintile (23 Dec 20 – 27 Dec 20)**

| **IMD Quintile** | | **Car Sharing or Taxi** | **Public Transport** | **Work or Education** | **Social/Entertainment** | **Essential Shops** | **Non-Essential Shops and Services** | **Healthcare Setting** | **Non-Household Contacts** |
|---|---|---|---|---|---|---|---|---|---|
| | total *n* | Yes % (*n*) | Yes % (*n*) | Yes % (*n*) | Yes % (*n*) | Yes % (*n*) | Yes % (*n*) | Yes % (*n*) | Yes % (*n*) |
| **1 (Most Deprived)** | 1630 | 16.20 (264) | 12.33 (201) | 22.39 (365) | 2.33 (38) | 67.85 (1106) | 11.66 (190) | 16.26 (265) | 40.92 (667) |
| **2** | 2958 | 12.07 (357) | 8.92 (264) | 21.20 (627) | 1.52 (45) | 67.04 (1983) | 10.18 (301) | 14.91 (441) | 37.19 (1100) |
| **3** | 4090 | 10.76 (440) | 5.79 (237) | 18.90 (773) | 2.30 (94) | 63.28 (2588) | 10.73 (439) | 14.91 (610) | 34.38 (1406) |
| **4** | 5262 | 10.49 (552) | 5.02 (264) | 18.26 (961) | 1.94 (102) | 64.61 (3400) | 10.70 (563) | 15.18 (799) | 33.69 (1773) |
| **5 (Least Deprived)** | 6180 | 8.72 (539) | 2.83 (175) | 16.49 (1019) | 1.68 (104) | 63.40 (3918) | 11.02 (681) | 14.63 (904) | 33.03 (2041) |
| **Total** | 20120 | 10.70 (2152) | 5.67 (1141) | 18.61 (3745) | 1.90 (383) | 64.59 (12995) | 10.81 (2174) | 15.00 (3019) | 34.73 (6987) |

**Note:** IMD = Indices of Multiple Deprivation

***Supplementary Table 3.***

**Proportion of participants reporting each activity and total number of respondents by deprivation quintile (09 Feb 21 – 16 Feb 21)**

| **IMD Quintile** | | **Car Sharing** | **Taxi** | **Public Transport** | **Work or Education** | **Social/Entertainment** | **Essential Shops** | **Non-Essential Shops and Services** | **Healthcare Setting** | **Non-Household Contacts** |
|---|---|---|---|---|---|---|---|---|---|---|
| | total *n* | Yes % (*n*) | Yes % (*n*) | Yes % (*n*) | Yes % (*n*) | Yes % (*n*) | Yes % (*n*) | Yes % (*n*) | Yes % (*n*) | Yes % (*n*) |
| **1 (Most Deprived)** | 2011 | 9.70 (195) | 5.72 (115) | 11.93 (240) | 25.21 (507) | 0.99 (20) | 66.53 (1338) | 6.46 (130) | 22.18 (446) | 40.13 (807) |
| **2** | 3780 | 9.44 (357) | 3.33 (126) | 10.16 (384) | 23.60 (892) | 0.48 (18) | 63.97 (2418) | 5.95 (225) | 21.61 (817) | 37.83 (1430) |
| **3** | 5174 | 8.95 (463) | 2.03 (105) | 5.68 (294) | 20.00 (1035) | 0.43 (22) | 61.77 (3196) | 5.51 (285) | 24.08 (1246) | 35.52 (1838) |
| **4** | 6629 | 8.33 (552) | 1.48 (98) | 4.22 (280) | 19.79 (1312) | 0.69 (46) | 62.50 (4143) | 6.59 (437) | 23.50 (1558) | 33.50 (2221) |
| **5 (Least Deprived)** | 7634 | 8.45 (645) | 0.83 (63) | 2.67 (204) | 18.77 (1433) | 0.45 (34) | 60.61 (4627) | 5.86 (447) | 23.08 (1762) | 34.35 (2622) |
| **Total** | 25228 | 8.77 (2212) | 2.01 (507) | 5.56 (1402) | 20.53 (5179) | 0.55 (140) | 62.32 (15722) | 6.04 (1524) | 23.11 (5829) | 35.35 (8918) |

**Note:** IMD = Indices of Multiple Deprivation

***Supplementary Table 4.***

**Risk Ratios for Work and Education by IMD Quintile for Children (<16 years) versus Adults (≥16 years)**

| **IMD Quintile** | **24 Nov 20 – 01 Dec 20** | | **23 Dec 20 – 27 Dec 20** | | **09 Feb 21 – 16 Feb 21** | |
|---|---|---|---|---|---|---|
| | **Child** | **Adult** | **Child** | **Adult** | **Child** | **Adult** |
| **1** | 0.96 [0.89,1.04] | ***1.30**** ***[1.18,1.43]*** | 0.53 [0.31,0.91] | ***1.41**** ***[1.25,1.59]*** | 0.90 [0.62,1.30] | ***1.39**** ***[1.26,1.55]*** |
| **2** | 0.94 [0.87,1.01] | ***1.28**** ***[1.18,1.39]*** | 0.90 [0.59,1.36] | ***1.31**** ***[1.18,1.45]*** | 0.71 [0.50,1.00] | ***1.33**** ***[1.22,1.45]*** |
| **3** | 0.98 [0.92,1.04] | 1.13 [1.04,1.22] | 0.67 [0.44,1.01] | ***1.21**** ***[1.09,1.34]*** | 0.66 [0.48,0.92] | 1.13 [1.04,1.23] |
| **4** | 1.03 [0.98,1.08] | 1.09 [1.01,1.17] | 0.87 [0.61,1.25] | 1.12 [1.02,1.23] | 0.86 [0.66,1.14] | 1.08 [1.00,1.17] |
| **5** | REF | REF | REF | REF | REF | REF |

**Note:** IMD = Indices of Multiple Deprivation (1 = most deprived; 5 = least deprived); 95% confidence intervals in parentheses
**** Bonferroni-Hochberg adjusted p<0.05***

*Supplementary Table 5.*

**Risk Ratios for Non-Household Contacts by IMD Quintile for Children versus Adults**

| **IMD Quintile** | **24 Nov 20 – 01 Dec 20** | | **23 Dec 20 – 27 Dec 20** | | **09 Feb 21 – 16 Feb 21** | |
|---|---|---|---|---|---|---|
| | **Child** | **Adult** | **Child** | **Adult** | **Child** | **Adult** |
| **1** | 0.92 [0.85,1.01] | ***1.11* [1.06,1.18]*** | 0.76 [0.54,1.08] | ***1.27* [1.17,1.38]*** | 0.93 [0.72,1.21] | ***1.20* [1.11,1.29]*** |
| **2** | 0.92 [0.85,1.00] | ***1.07* [1.02,1.12]*** | 0.97 [0.74,1.27] | ***1.14* [1.06,1.23]*** | 0.91 [0.72,1.16] | ***1.12* [1.05,1.20]*** |
| **3** | ***0.91* [0.84,0.97]*** | 1.03 [0.99,1.08] | 0.70 [0.53,0.93] | 1.08 [1.01,1.16] | 0.73 [0.57,0.93] | 1.06 [1.00,1.13] |
| **4** | 0.99 [0.93,1.04] | 1.04 [1.00,1.09] | 0.75 [0.58,1.96] | 1.04 [0.98,1.11] | 0.96 [0.79,1.17] | 0.98 [0.93,1.04] |
| **5** | REF | REF | REF | REF | REF | REF |

**Note:** IMD = Indices of Multiple Deprivation (1 = most deprived; 5 = least deprived); 95% confidence intervals in parentheses
***** Bonferroni-Hochberg adjusted p<0.05***

*Supplementary Table 6.*

**Risk Ratios for Exposure to Work and Education Settings and Non-Household Contacts by IMD Quintile for Adults - Controlling for Presence of Child(ren) in the Household**

| IMD Quintile | 24 Nov 20 – 01 Dec 20 | | 23 Dec 20 – 27 Dec 20 | | 09 Feb 21 – 16 Feb 21 | |
|---|---|---|---|---|---|---|
| | **Work or Education** | **Non-Household Contacts** | **Work or Education** | **Non-Household Contacts** | **Work or Education** | **Non-Household Contacts** |
| **1** | ***1.28 [1.17, 1.41]**** | ***1.11 [1.05, 1.17]**** | ***1.38 [1.23, 1.56]**** | ***1.26 [1.16, 1.38]**** | ***1.38 [1.25, 1.53]**** | ***1.20 [1.11, 1.29]**** |
| **2** | ***1.29 [1.19, 1.39]**** | ***1.07 [1.02, 1.12]**** | ***1.31 [1.18, 1.45]**** | ***1.14 [1.06, 1.23]**** | ***1.34 [1.23, 1.46]**** | ***1.12 [1.05, 1.20]**** |
| **3** | ***1.14 [1.06, 1.23]**** | 1.03 [0.99, 1.08] | ***1.22 [1.10, 1.34]**** | ***1.08 [1.01, 1.16]**** | ***1.14 [1.05, 1.24]**** | ***1.06 [1.00, 1.13]**** |
| **4** | 1.09 [1.02, 1.17] | 1.05 [1.00, 1.09] | 1.13 [1.03,1.24] | 1.04 [0.98, 1.11] | 1.09 [1.01, 1.18] | 0.98 [0.93, 1.04] |
| **5** | REF | REF | REF | REF | REF | REF |

**Note:** IMD = Indices of Multiple Deprivation (1 = most deprived; 5 = least deprived); 95% confidence intervals in parentheses
**** Bonferroni-Hochberg adjusted p<0.05***

*Supplementary Figure 1a.* **Risk Ratios for Public Activities and Non-Household Contacts by IMD Quintile (24 Nov 20 - 01 Dec 20)**

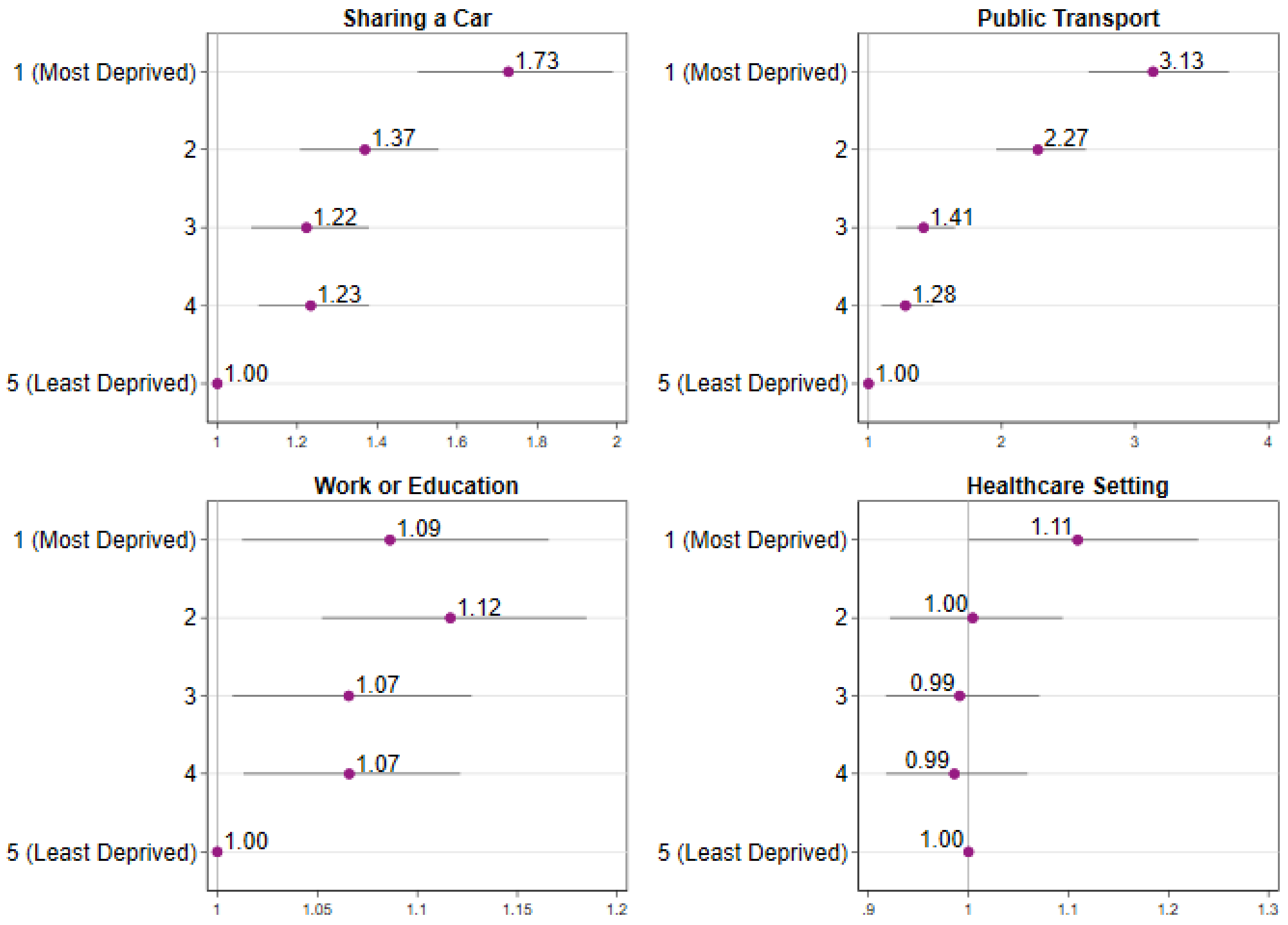

***Supplementary Figure 1b.*** **Risk Ratios for Public Activities and Non-Household Contacts by IMD Quintile (24 Nov 20 - 01 Dec 20)**

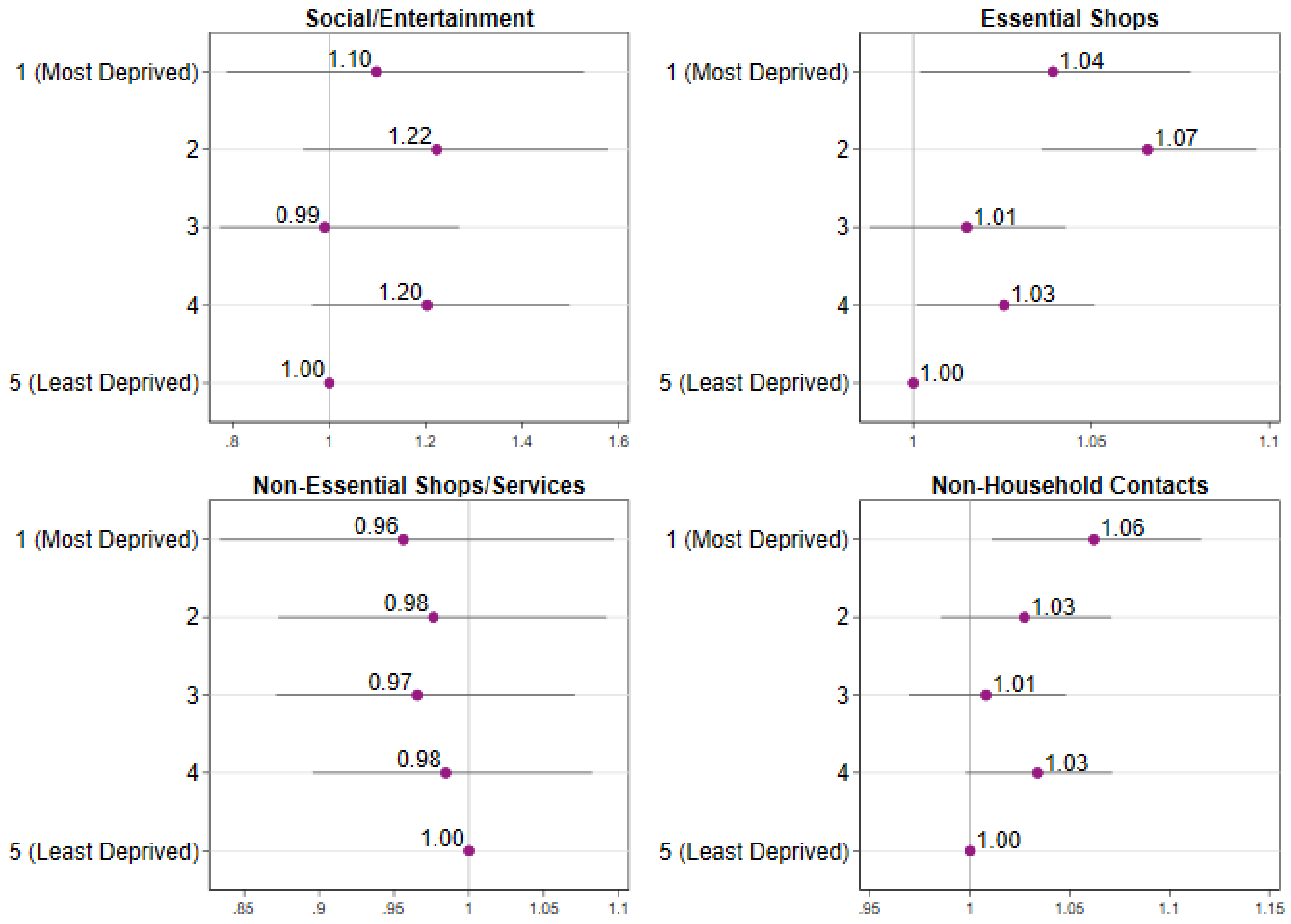

*Supplementary Figure 2a.* **Risk Ratios for Public Activities and Non-Household Contacts by IMD Quintile (23 Dec 20 - 27 Dec 20)**

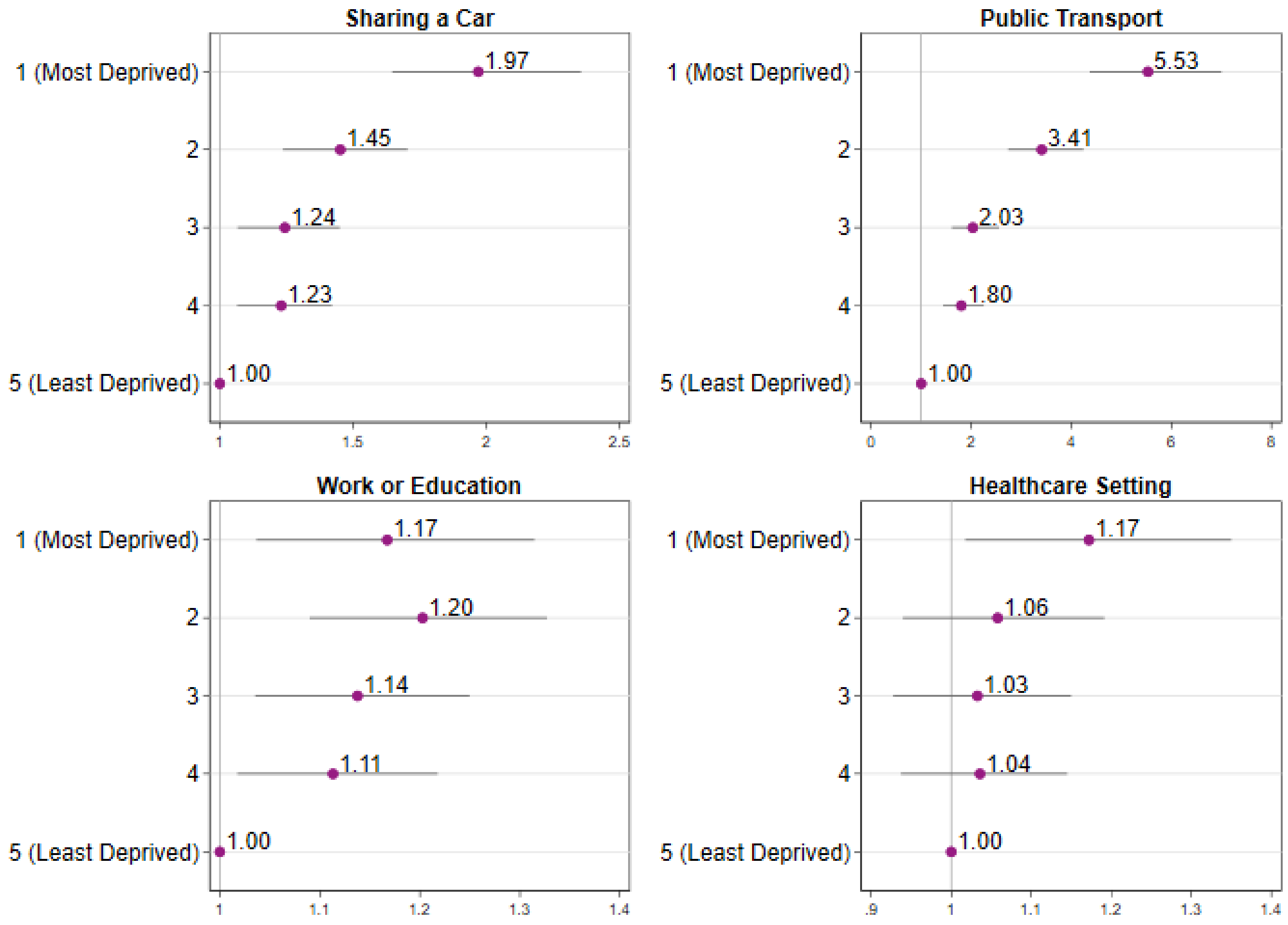

***Supplementary Figure 2b.*** **Risk Ratios for Public Activities and Non-Household Contacts by IMD Quintile (23 Dec 20 - 27 Dec 20)**

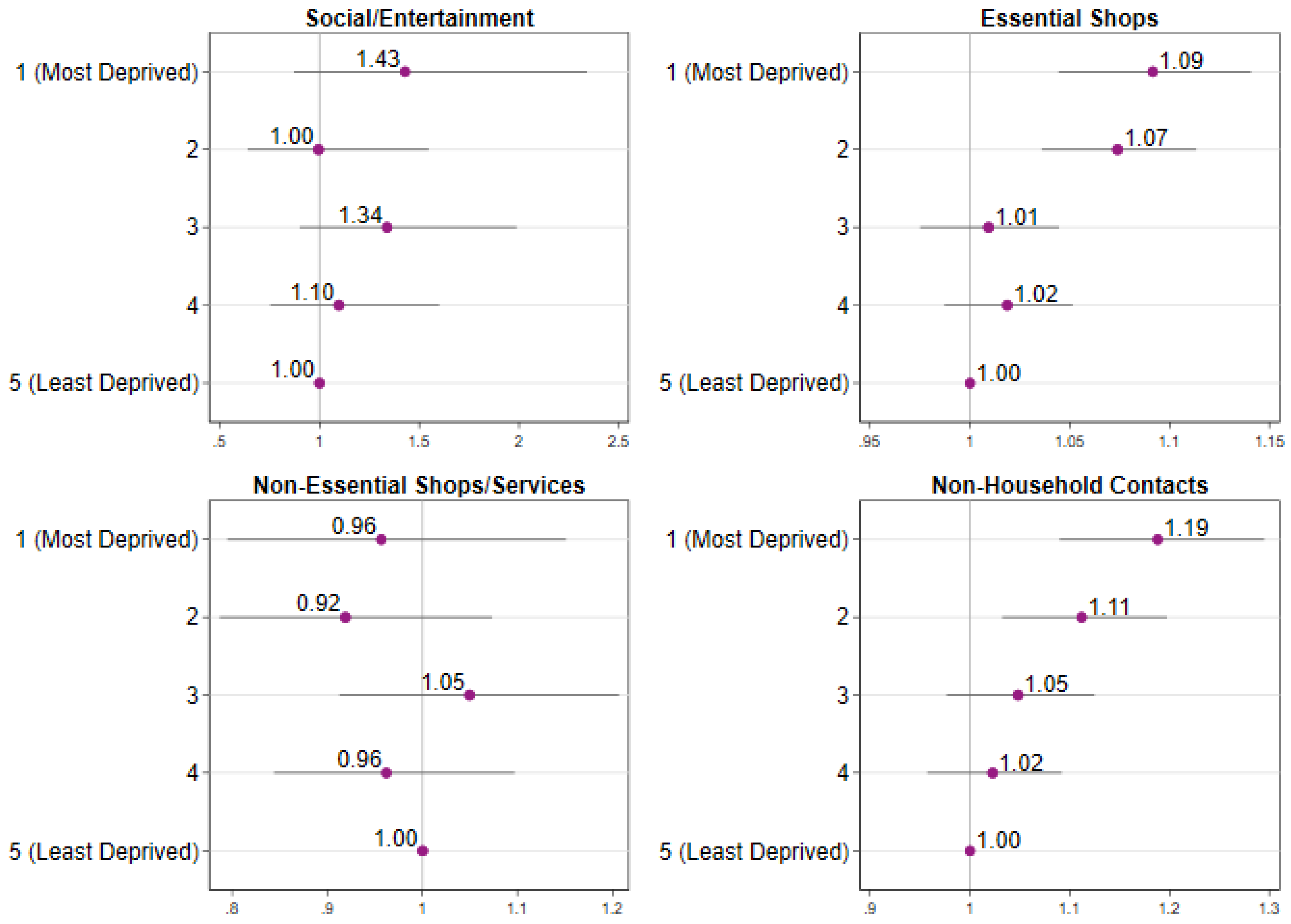

***Supplementary Figure 3a.*** **Risk Ratios for Public Activities and Non-Household Contacts by IMD Quintile (09 Feb 21 - 16 Feb 21)**

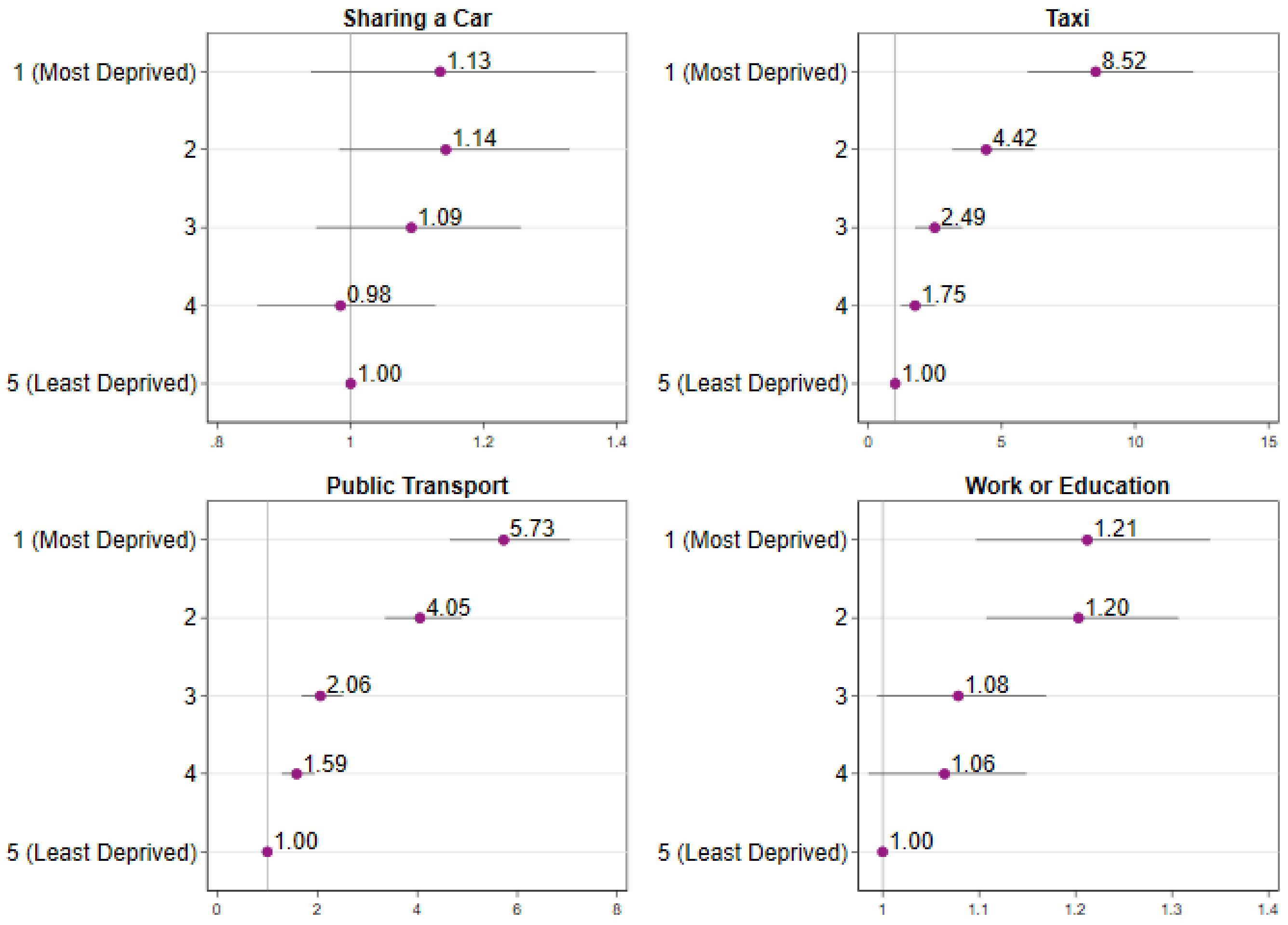

***Supplementary Figure 3b.*** **Risk Ratios for Public Activities and Non-Household Contacts by IMD Quintile (09 Feb 21 - 16 Feb 21**

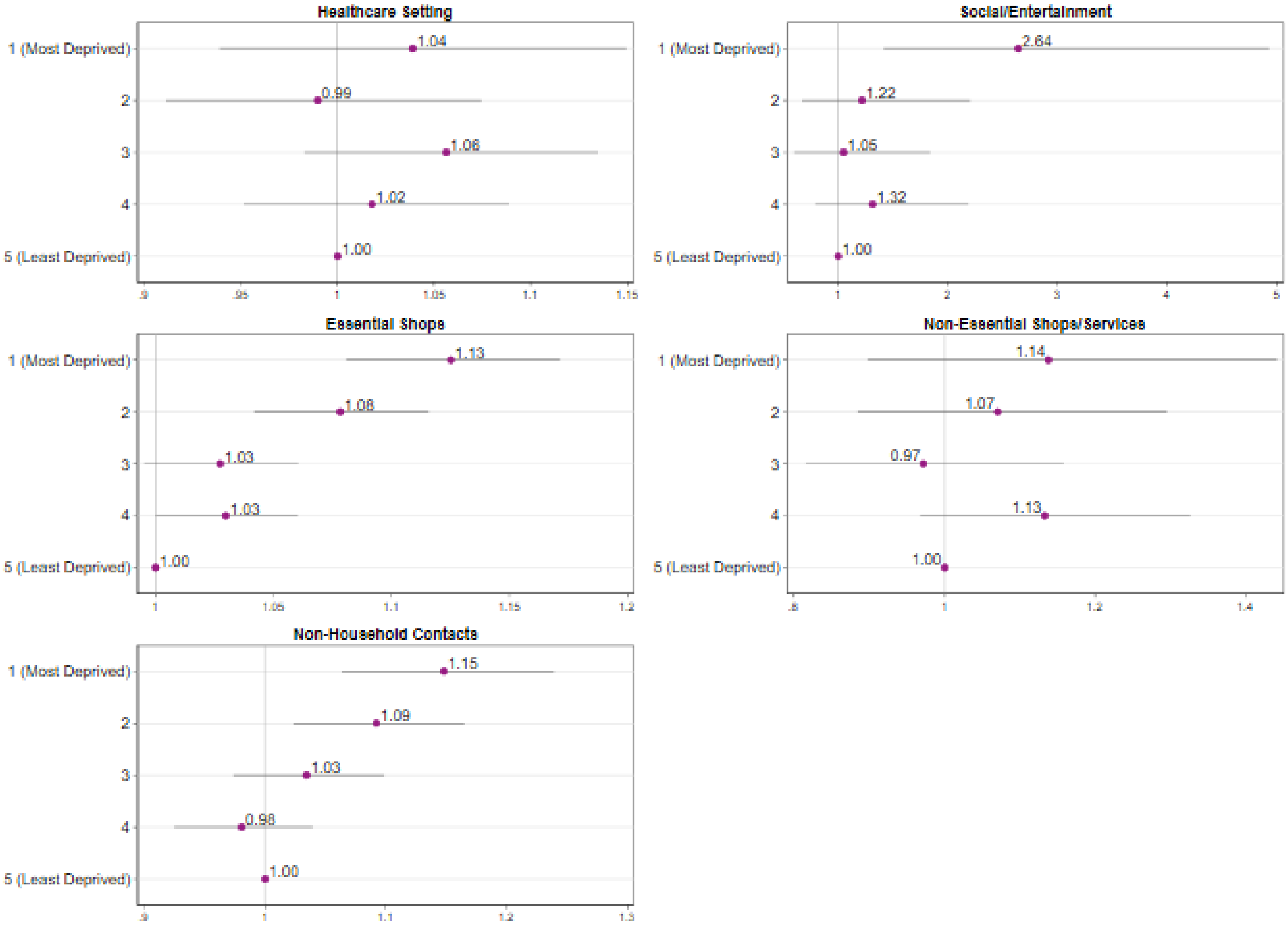
